# Characterizing Variability of EHR-Driven Phenotype Definitions

**DOI:** 10.1101/2022.07.10.22277390

**Authors:** Pascal S. Brandt, Abel Kho, Yuan Luo, Jennifer A. Pacheco, Theresa L. Walunas, Hakon Hakonarson, George Hripcsak, Cong Liu, Ning Shang, Chunhua Weng, Nephi Walton, David S. Carrell, Paul K. Crane, Eric Larson, Christopher G. Chute, Iftikhar Kullo, Robert Carroll, Josh Denny, Andrea Ramirez, Wei-Qi Wei, Jyoti Pathak, Laura K. Wiley, Rachel Richesson, Justin B. Starren, Luke V. Rasmussen

## Abstract

**Objective:** Analyze a publicly available sample of rule-based phenotype definitions to characterize and evaluate the types of logical constructs used.

**Materials & Methods:** A sample of 33 phenotype definitions used in research and published to the Phenotype KnowledgeBase (PheKB), that are represented using Fast Healthcare Interoperability Resources (FHIR) and Clinical Quality Language (CQL) was analyzed using automated analysis of the computable representation of the CQL libraries.

**Results:** Most of the phenotype definitions include narrative descriptions and flowcharts, while few provide pseudocode or executable artifacts. Most use 4 or fewer medical terminologies. The number of codes used ranges from 5 to 6865, and value sets from 1 to 19. We found the most common expressions used were literal, data, and logical expressions. Aggregate and arithmetic expressions are the least common. Expression depth ranges from 4 to 27.

**Discussion:** Despite the range of conditions, we found that all of the phenotype definitions consisted of logical criteria, representing both clinical and operational logic, and tabular data, consisting of codes from standard terminologies and keywords for natural language processing. The total number and variety of expressions is low, which may be to simplify implementation, or authors may limit complexity due to data availability constraints.

**Conclusion:** The phenotypes analyzed show significant variation in specific logical, arithmetic and other operators, but are all composed of the same high-level components, namely tabular data and logical expressions. A standard representation for phenotype definitions should support these formats and be modular to support localization and shared logic.

## BACKGROUND AND SIGNIFICANCE

The generation of biomedical knowledge from electronic health record (EHR) data requires establishing cohorts of patients meeting certain criteria.[Banda2018] This cohort identification process is referred to as *EHR-driven phenotyping* and the sets of inclusion and exclusion criteria are known as *phenotype definitions*, or just *phenotypes* (the term used here for brevity). Although here we focus on the use within a research context, phenotypes support a wide variety of purposes beyond research, including quality improvement, clinical decision support, population health management, and public health. While often developed and executed at a single institution, research networks such as the National Patient-Centered Clinical Research Network (PCORnet),[Fleurence2014] the electronic Medical Records and Genomics (eMERGE) Network, [McCarty2011, Gottesman2013, Zouk2019] and the Observational Health Data Sciences and Informatics (OHDSI) program [Hripcsak2015] have run distributed studies to pool their results to improve statistical power and cohort diversity, leveraging shared phenotype definitions to meet these goals.[Ahmad2020, Hripcsak2019, Burn2020]

Beyond coordinated research networks, the re-use of phenotype definitions can save time across organizations and create many efficiencies.[Richesson2016] But this requires that potential users be able to search, retrieve, and assess existing definitions. At present, there is no widely used platform for sharing existing phenotype definitions, although multiple phenotype libraries or inventories have been created within research consortia. Historically, phenotypes have been shared between sites via repositories in the form of narrative descriptions,[Kirby2016, Denaxas2019] sometimes accompanied by flowcharts or pseudocode. Lists of codes from common terminologies like the International Classification of Diseases version 9 (ICD-9) are usually also included, but in most cases directly computable artifacts, such as SQL scripts or programming code, are not. Despite the potential gains of phenotype re-use, this current method of phenotype distribution has proven to be a major limiting factor in scaling up biomedical knowledge generation,[Pathak2013a, Newton2013, Shivade2014a, Adekkanattu2020] since implementing sites must individually interpret narrative descriptions to produce queries that can extract patient cohorts from local data sources. This process is time-consuming, may be operator dependent and error prone, compounded by the fact that narrative descriptions can be ambiguous or difficult to interpret.[Yu2021]

However, at present there is no universally accepted standard for representing the attributes and specification for computable phenotype definitions, although several have been proposed and evaluated [Peterson2014a, Pathak2013, Mo2015b, Mo2016, Jiang2017, Chapman2020a, Hong2019, Brandt2020]. The lack of a common representation for computable phenotypes has hindered the analysis and comparison of different phenotype algorithms. For example, it is not possible to do an equal technical comparison of a phenotype implemented in SQL against a local data warehouse and one done in Python against the OMOP CDM. Previous analyses have been performed in a few domains including an early review of 11 narrative phenotype descriptions;[Conway2011] clinical quality measures (CQMs);[Dorr2011] and high-level categorizations of the elements that make up clinical trial inclusion and exclusion criteria.[VanSpall2007, Ross2010a] Comparisons of multiple phenotype definitions have also been done,[Richesson2013] but have focused on a single disease or condition. However, no work to date has evaluated phenotype composition across diseases using a consistent computable representation.

The goal of this study is to characterize EHR-based phenotype definitions using a consistent computable representation, and generate new knowledge about the types and variability of constructs that must be accommodated in a system that is capable of formally representing a rich and diverse set of phenotype algorithms.

## Materials & Methods

### Data Set

We used a set of 33 phenotype definitions that were represented using FHIR and CQL. Full details about the creation of the phenotype definitions are explained elsewhere,[Brandt2021] but briefly, the data set is comprised of phenotype algorithms that utilized structured data, were marked as “Final” in PheKB, and were used in a published research study. These phenotype algorithms were then translated into FHIR (selected for its growth and use in healthcare and research contexts) and CQL (selected for its use within eCQMs and ability to represent phenotype algorithms), and validated using manual review and automated testing. The dataset is open source and available on GitHub^1^.

### Data Analysis

#### Metadata Analysis

PheKB allows phenotype submitters to voluntarily annotate their phenotype algorithm with metadata. We extracted and reviewed the metadata available on the PheKB page for each phenotype in JSON format, but this metadata was not used in our analysis due to issues with irrelevant, incomplete or outdated information. Instead, supplemental metadata was manually assembled by two authors (PSB, LVR) who independently conducted a review of PheKB for each phenotype in the dataset and curated relevant dimensions, emergent patterns, and characteristics. We categorized the artifacts provided with each phenotype definition (e.g., flowcharts) and whether or not the definition for controls, subtypes, or suspected cases is provided. We also provided a “Type” categorization to capture the intent of the phenotype, and note whether or not the phenotype used tabular data, and how these data are provided. In most cases tabular data refers to lists of codes from standard terminologies, but may also include lists of keywords or medication names. Following this manual review, the two reviewers met to discuss their findings and resolved any discrepancies.

#### Phenotype Definition Analysis

To evaluate the phenotype definition logic, we conducted an automated analysis of the Expression Logical Model (ELM) representation of each CQL library. An ELM representation is an instance of an Abstract Syntax Tree (AST),[Parr2009] which acts as a machine-readable representation of a complete program and is used to evaluate or execute the program. ASTs can be used in program translation, as has been shown for CQL,[Brandt2020] or for program analysis, as we demonstrate here. We evaluated the ELM for each phenotype by making use of the Visitor Pattern,[Gamma1995] which is a mechanism for inspecting each node of tree-like data structures and executing custom code in the context of each node. Our Java implementation used an interface provided by the reference implementation of the CQL translator,^2^ which is the same one used by the CQL engine during program execution. Our evaluator program calculated a number of measures about a given CQL library, including how many value sets are referenced, how many Boolean, temporal, and aggregate operators are used, and how these operators are combined. We also determined the total number of expressions, how many data types are used, and how many unique data queries are performed. After a preliminary manual review of the phenotype definitions, we identified 11 dimensions along which to evaluate each phenotype, shown in Table 1. We note that the library used did not include NLP implementations, and so NLP-related metrics were not considered in our analysis.

**Table 1.** Phenotype definition analysis dimensions.

| Category | Description | Examples |
| --- | --- | --- |
| Aggregate | Operations that calculate single values from collections | **sum**, **count** or **mean** |
| Arithmetic | Mathematical operations | +, -, or * |
| Collection | Operations on collections of data like sets and lists | **first**, **exists** or **union** |
| Comparison | Numeric or date comparisons | > or = |
| Conditional | Branching logic | **if** or **case** |
| Data | Data retrieval and filtering operations | FHIR resource querying and filtering by value set |
| Expressions | Total number of expressions as well as their depth | Total expression count, where clause expression depth |
| Literals | Explicit values, codes and quantities | **23**, **5 months** or **0.5 mg/dL** |
| Logical | Boolean logical operators | **and** or **not** |
| Temporal | Operators relating to dates and times | **before**, **starts** or **overlaps** |
| Terminology | Number of value sets used and the number of individual codes | Value sets per phenotype, codes per code system |

## Results

### Metadata

Table 2 provides metadata extracted by manually reviewing each phenotype definition. Our manually determined types align with the self-reported PheKB types, but we introduce a new type with the label “Valid Data”. This indicates that the phenotype is trying to identify patients without disqualifying data. For example, the *Height* phenotype identifies individuals who have a valid height measurement and do not have any conditions that may impact height. We also introduce the “Treatment / Therapy” type, which identifies individuals who have had a specific treatment, for example, *Bone Scan Utilization*. Finally, we describe the approach outlined for NLP implementation (if applicable).

**Table 2.** Metadata manually extracted from PheKB. **NC** – Non-computable (e.g., Word or PDF), **XLSX** – Microsoft Excel file, **CSV** – Comma Separated Value file, **KNIME** – KoNstanz Information MinEr files, **SQL** – Structured Query Language

| Name | Type | Narrative | Flowchart | Pseudocode | Tabular | Executable | Suspected | Controls | Subtypes | Covariates | NLP |
| --- | --- | --- | --- | --- | --- | --- | --- | --- | --- | --- | --- |
| Asthma Response to Inhaled Steroids | Drug Response | ✓ |  |  | NC |  |  |  |  |  | regex |
| Atrial Fibrillation | Disease |  |  | ✓ | NC |  | ✓ | ✓ | ✓ |  | keywords; regex |
| Autism | Disease | ✓ | ✓ |  | NC |  |  | ✓ | ✓ | ✓ | DSM-IV criteria |
| Benign Prostatic Hyperplasia | Disease |  | ✓ | ✓ | NC | KNIME |  | ✓ |  | ✓ | keywords |
| Bone Scan Utilization | Treatment / Therapy | ✓ | ✓ |  | NC |  |  | ✓ |  |  | keywords |
| Cardiac Conduction | Trait | ✓ |  |  | NC |  |  |  |  | ✓ | keywords; negation; uncertainty |
| Cataracts | Disease |  | ✓ | ✓ | NC |  |  | ✓ | ✓ | ✓ | MedLEE concepts; negation; regex |
| Clopidogrel Poor Metabolizers | Drug Response |  | ✓ |  | NC |  |  | ✓ | ✓ |  | keywords |
| Crohn's Disease | Disease |  |  | ✓ | NC |  | ✓ | ✓ | ✓ |  | keywords; regex |
| Developmental Language Disorder | Disease | ✓ | ✓ |  | CSV | KNIME |  |  | ✓ |  |  |
| Digital Rectal Exam | Treatment / Therapy | ✓ | ✓ |  | NC |  |  | ✓ |  |  | documentation is obtained from clinical notes |
| Drug Induced Liver Injury | Drug Response | ✓ | ✓ |  | NC |  |  | ✓ |  | ✓ | medication names; "diagnosis mentioned" |
| Familial Hypercholesterolemia | Disease | ✓ | ✓ | ✓ | XLSX |  |  | ✓ |  | ✓ | detailed pseudocode |
| Height | Healthy / Valid Data |  | ✓ |  | NC | KNIME |  |  |  | ✓ | keywords |
| Herpes Zoster | Disease |  | ✓ | ✓ | XLSX |  |  | ✓ |  | ✓ |  |
| High-Density Lipoproteins | Trait |  | ✓ | ✓ | NC |  |  |  |  |  | keywords |
| Hypothyroidism | Disease | ✓ |  |  | NC |  |  | ✓ |  | ✓ | keywords |
| Lipids | Healthy / Valid Data | ✓ | ✓ | ✓ | NC | KNIME |  |  |  | ✓ |  |
| Multimodal Analgesia | Treatment / Therapy | ✓ | ✓ |  | NC |  |  | ✓ |  |  | medication names |
| Multiple Sclerosis | Disease |  |  | ✓ | NC |  | ✓ | ✓ |  |  | keywords; regex |
| Peripheral Arterial Disease | Disease | ✓ |  |  | NC |  | ✓ |  |  | ✓ | keywords |
| Red Blood Cell Indices | Healthy / Valid Data | ✓ | ✓ |  | NC |  |  |  |  | ✓ | medication names |
| Resistant hypertension | Drug Response | ✓ |  |  | NC |  |  | ✓ |  | ✓ | medication names |
| Rheumatoid Arthritis | Disease |  |  | ✓ | NC |  | ✓ | ✓ |  |  | keywords; regex |
| Sickle Cell Disease | Disease | ✓ |  |  | NC |  |  |  |  |  |  |
| Statins and MACE | Drug Response | ✓ | ✓ |  | NC |  |  | ✓ | ✓ | ✓ | medication names; keywords |
| Steroid Induced Osteonecrosis | Drug Response |  | ✓ |  | NC |  |  | ✓ |  |  | keywords |
| Systemic Lupus | Disease | ✓ |  |  |  |  |  |  |  |  | keywords |
| Type 2 Diabetes | Disease |  |  | ✓ | NC |  |  | ✓ |  |  | keywords; regex |
| Type 2 Diabetes Mellitus | Disease | ✓ | ✓ | ✓ | XLSX | KNIME; SQL fragments |  | ✓ |  | ✓ | medication names; regex |
| Urinary Incontinence | Treatment / Therapy | ✓ |  |  |  |  |  |  |  |  | executable Python code |
| Warfarin Dose/Response | Drug Response |  | ✓ |  |  |  |  |  | ✓ |  | keywords |
| White Blood Cell Indices | Healthy / Valid Data | ✓ |  |  | NC |  |  |  |  | ✓ |  |

About two thirds of the definitions provide a narrative description (n=20) and flowchart (n=19), while only about one third (n=12) provide pseudocode. While tabular data is provided by all but three phenotypes, only 4 provide this data in a computable format. Computable artifacts in the form of KNIME workflows are provided for 5 phenotypes, and we found that these workflows require users to prepare their data in a specified custom format before execution.

Most phenotypes (n=20) provide control definitions, 8 provide phenotype subtype definitions, and 4 include a category for suspected cases. About half (n=16) of the phenotypes provide a list of covariates to be collected. All but 5 phenotypes rely on some form of NLP, with 17 providing a list of keywords, 8 providing regular expressions, and 6 providing a list of medication names.

### Terminologies

Figures 2 and 3 provide histograms related to codes, code systems and value. The figures were generated using automated analysis of the ELM representation of the phenotype definitions and the value sets in FHIR format. Most phenotypes (n=28) use four or fewer code systems, and almost all (n=30) use ICD-9 codes, driven in part by the prevalence of phenotype algorithms in PheKB developed prior to or shortly after the adoption of ICD-10 in the United States. RxNorm (n=21), LOINC® (n=17), and CPT (n=16) are the next most commonly used. Five code systems (AMT, dm+d, BDPM, CIEL, and MedDRA) are each only used by a single phenotype.

**Figure 2.**
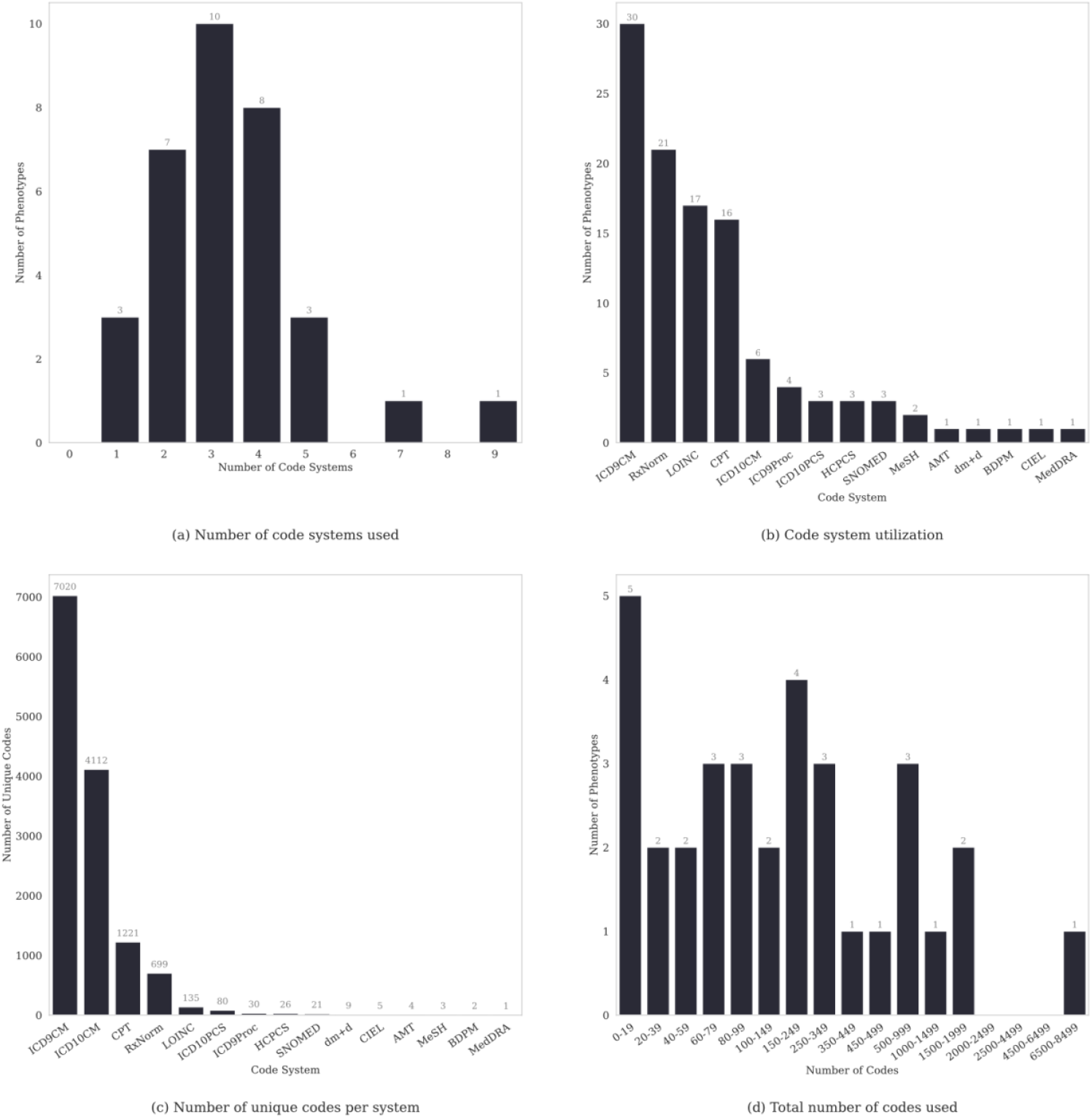
Histograms of code and code system usage. **ICD9CM** – International Classification of Diseases, Ninth Revision, Clinical Modification; **ICD10CM** – International Classification of Diseases, Tenth Revision, Clinical Modification; **LOINC** - Logical Observation Identifiers Names and Codes; **CPT** - Current Procedural Terminology; **ICD9Proc** – International Classification of Diseases, Ninth Revision, Procedures; ; **ICD10PCS** – International Classification of Diseases, Tenth Revision, Procedure Coding System; **HCPCS** - Healthcare Common Procedure Coding System; **SNOMED** – Systematized Nomenclature of Medicine; **MeSH** - Medical Subject Headings; **AMT** - Australian Medicines Terminology; **dm+d** - Dictionary of Medicines and Devices; **BDPM** - Public Database of Medications; **CIEL** - Columbia International eHealth Laboratory; **MedDRA** - Medical Dictionary for Regulatory Activities

**Figure 3.**
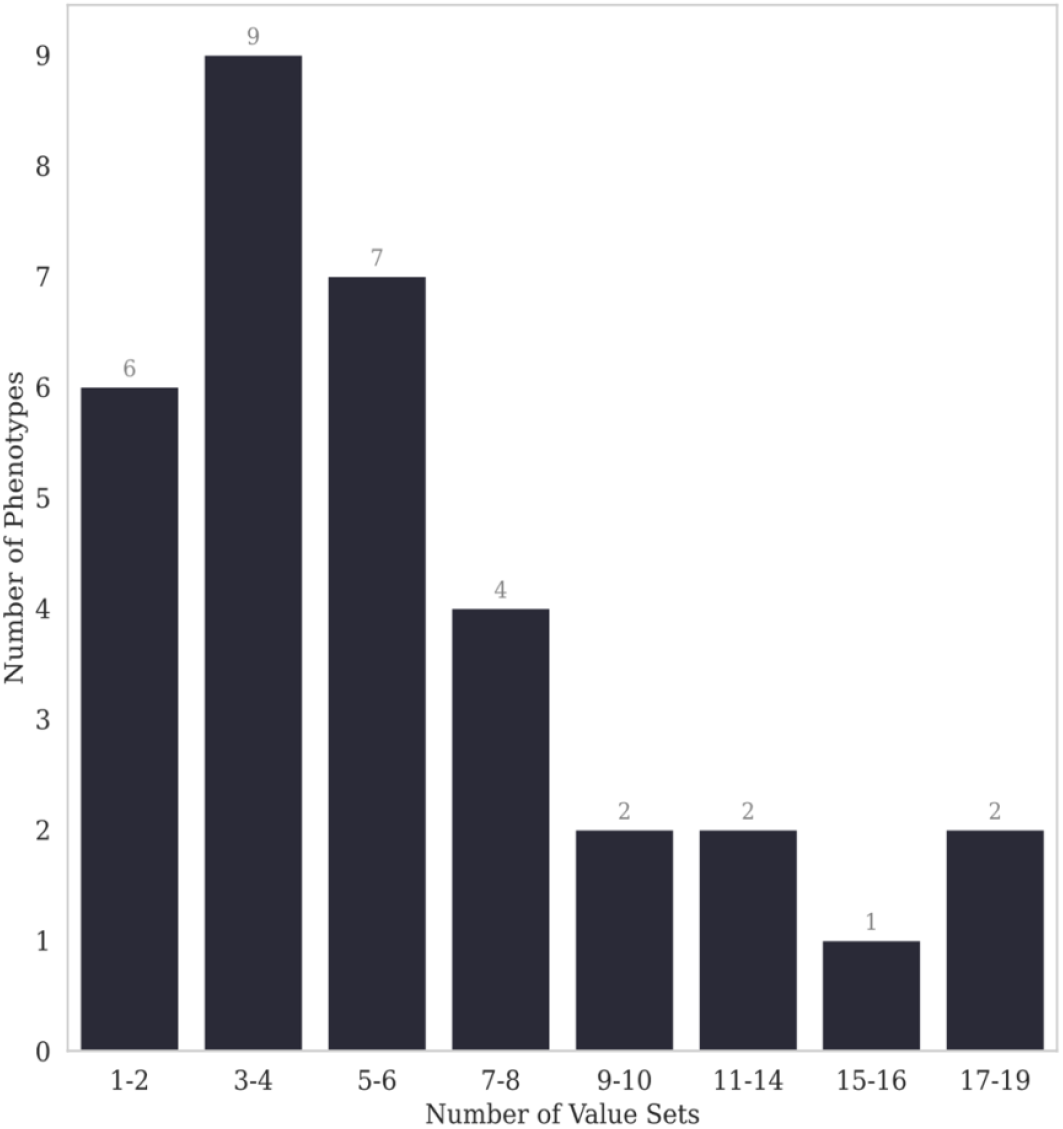
Histogram of value sets used.

About half of all codes (n=7020) are ICD-9 codes, and about a quarter (n=4112) are ICD-10 codes. CPT (n=1221) and RxNorm (n=699) are the next most common. The total number of codes used varies from 5 (*Warfarin Dose/Response*) to 6865 (*Developmental Language Disorder*), with a median of 147 (mean: 509.2, std: 1206.3). The total number of value sets used ranges from 1 (*Lipids and Sickle Cell Disease*) to 19 (*Resistant Hypertension*), with a median of 5 (mean: 6, std: 4.4).

### Logical Expressions

Logical expressions were analyzed by programmatically examining the ELM representation of each phenotype definition. Figure 4 illustrates the total number of expressions used in each phenotype broken down by CQL expression category, enabling easy comparison between phenotypes and demonstrating the range of variability among the definitions. Figure 5 provides a histogram of the individual expressions within each category and Figure 6 illustrates the data types utilized by literal expressions.

**Figure 4.**
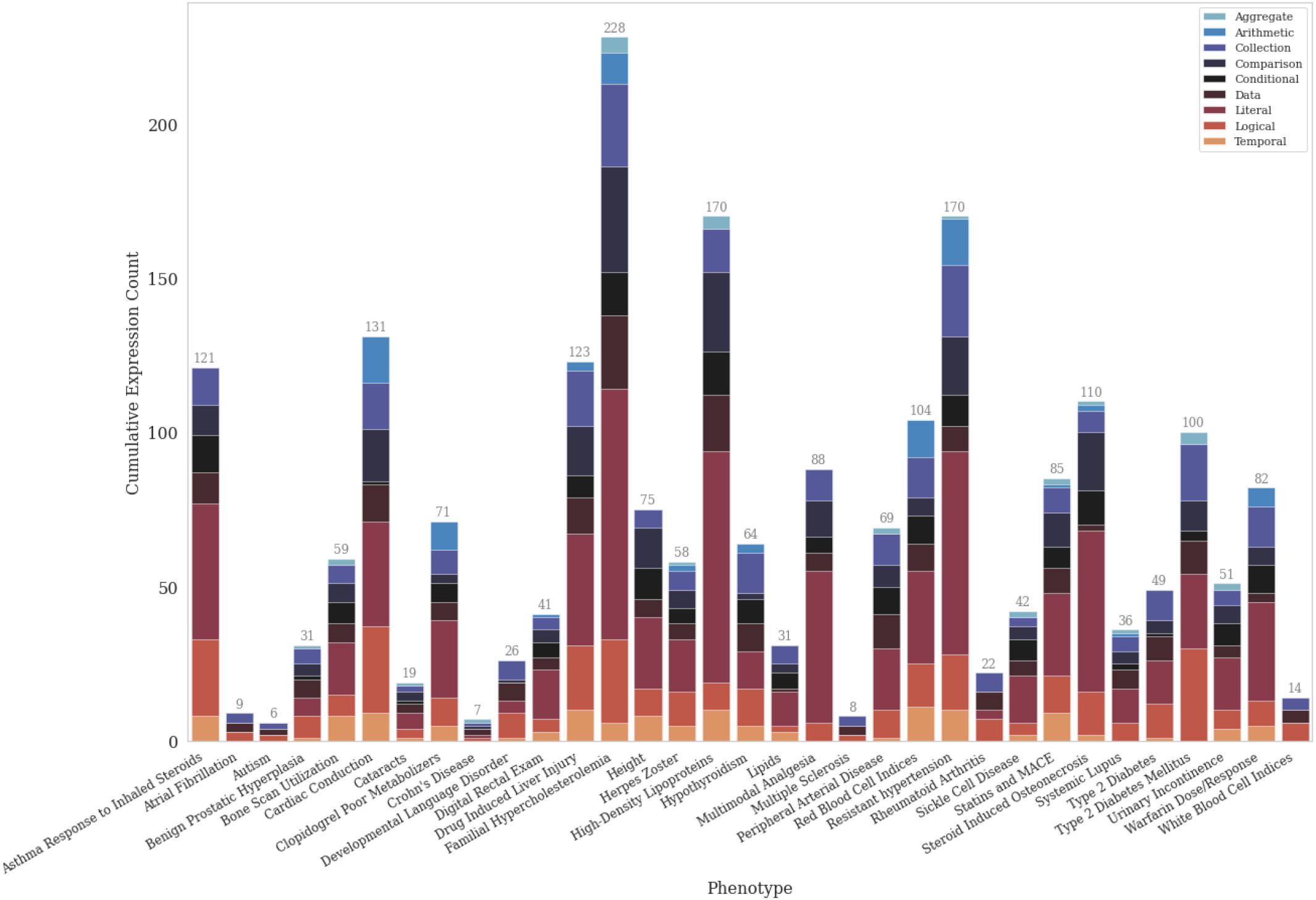
Cumulative expression counts per category.

**Figure 5.**
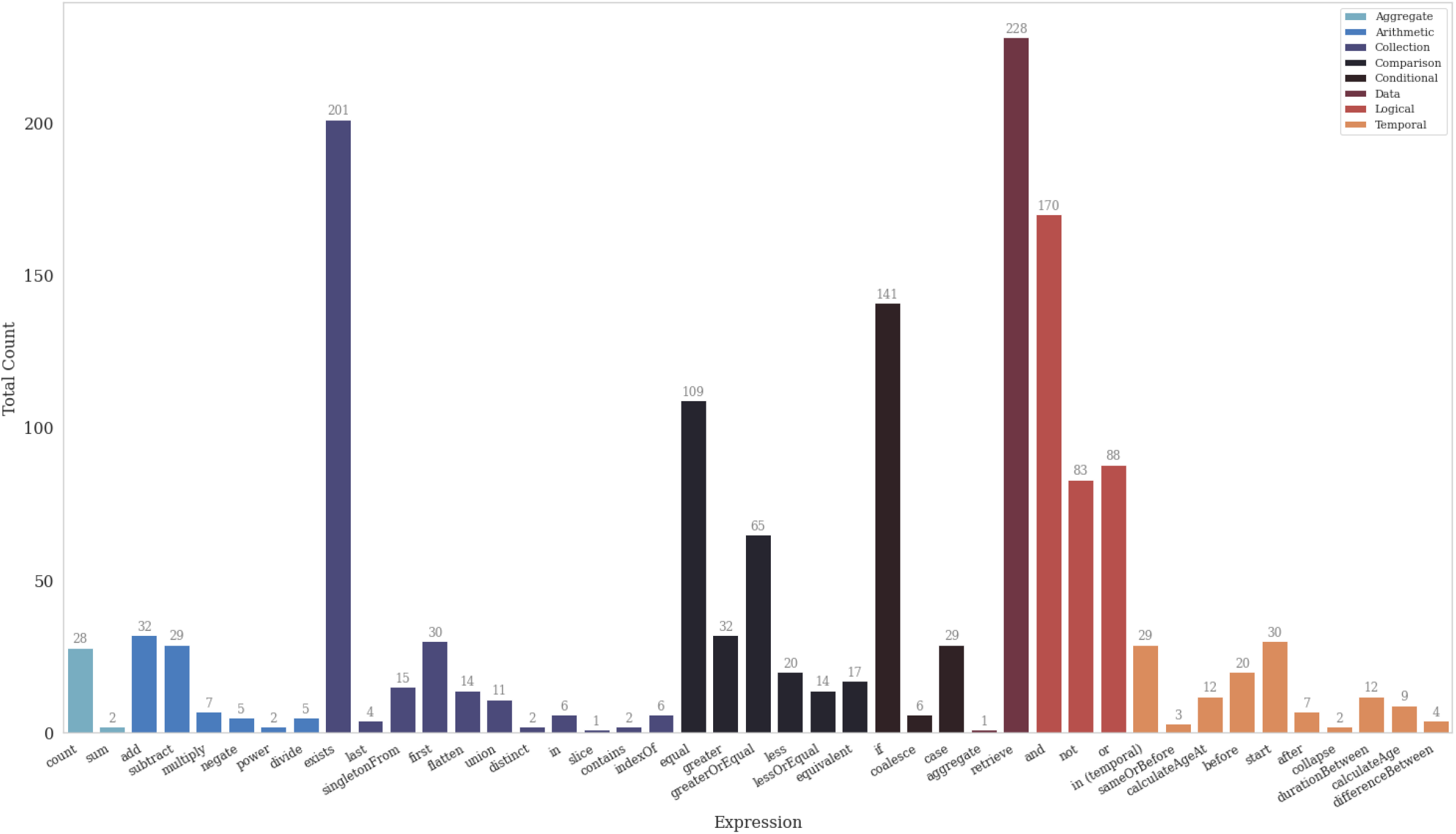
Total numbers of individual expression types by category (excluding literal expressions).

**Figure 6.**
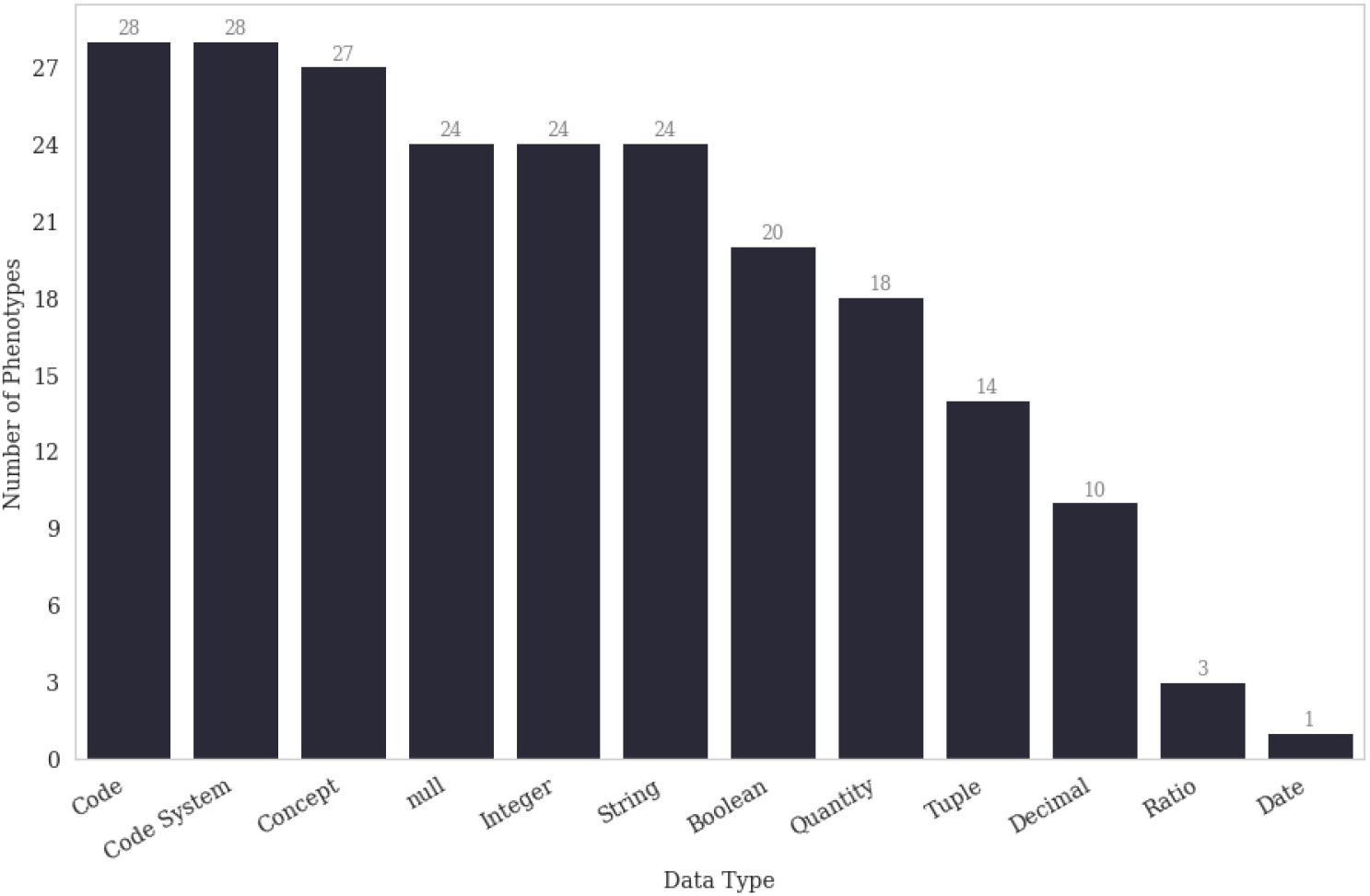
Utilization of various data types by literal expressions.

The most widely used expression categories are literal (n=767), data (n=229), logical (n=341), and collection (n=292), with the latter three categories used by every phenotype. The least commonly used expression categories are aggregate (n=30) and arithmetic (n=80). The total number of expressions used ranges from 6 (*Autism*) to 228 (*Familial Hypercholesterolemia*), with a median of 59 (mean: 69.7, std: 53.3).

Only two types of aggregate expressions were used, with `count` (n=28) being the most common. The `exists` (n=191) expression is the most common collection expression used, with `equal` (n=109) and `if` (n=141) the most common comparison and conditional expressions respectively. The `retrieve` (n=228) expression was the most common non-literal expression overall, while the `aggregate` data expression was used only once. The `and` (n=170) expression was the most common logical operator, being used about twice as many times as `not` (n=83) and `or` (n=88). The `start` (n=30), which extracts the start date or time from an interval, is the most common temporal operator.

Figure 6 shows the frequency of data types for literal expressions. Of these, terminology literals are the most commonly used types, with the `code`, `code system`, and `concept` types each occurring in 27 or more phenotypes. The next most common are primitive types like `Boolean` (20) and `Integer` (24). In total, 18 phenotypes make use of a `Quantity` data type (a scalar value with a unit).

### Data Sources

Figure 7 illustrates how many data types (distinct FHIR resources, such as Condition or Encounter, given the use of FHIR as the data model) are used per phenotype definition and how many phenotypes used each data source. The majority of phenotypes (n=22) used 3 data types or fewer. Condition was the most common data type, used by almost all (n=30) phenotypes, followed by medications (n=22), procedures (n=17), and observations (n=17). Demographic and encounter data were the least frequently used.

**Figure 7.**
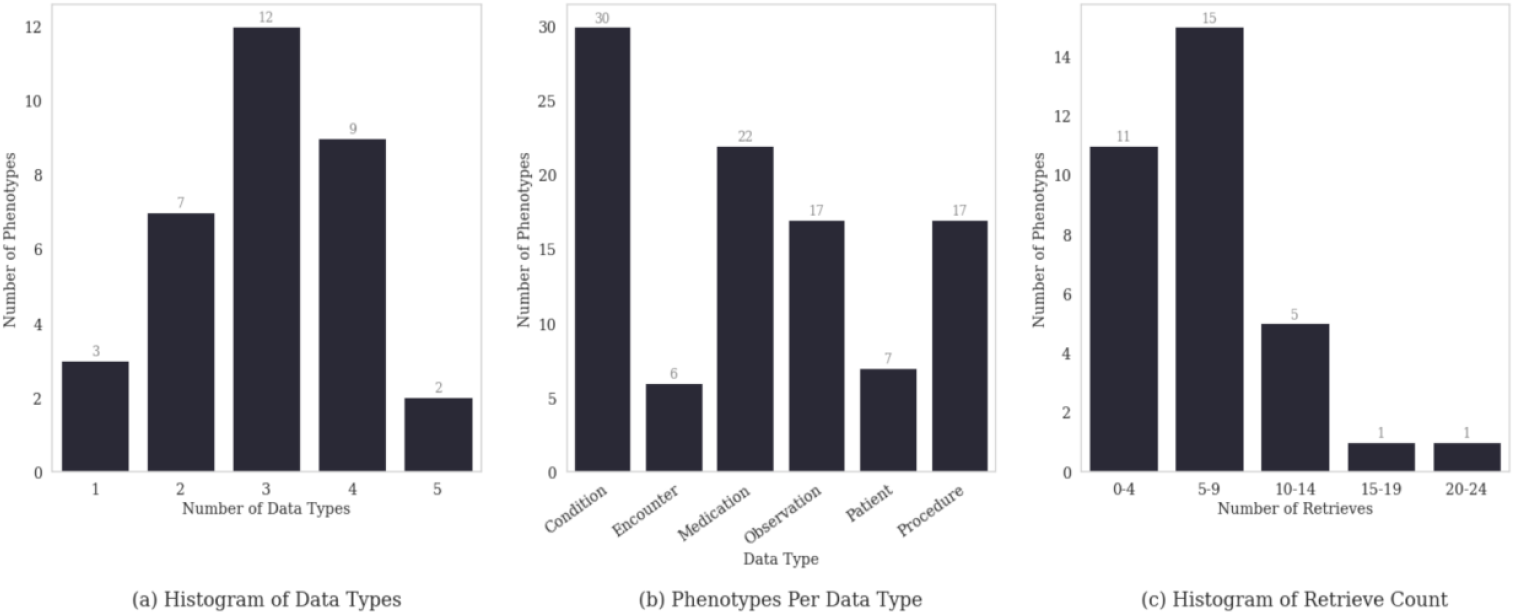
Data retrieval expressions and data types.

Also provided are histograms of the numbers of `retrieve` operations (used to fetch records for data sources). The majority of phenotypes use fewer than ten retrieves. The outliers are *Familial Hypercholesterolemia* (n=24) and *High-Density Lipoproteins* (n=18). The number of retrieves vary depending on the phenotype logic. For example, *Resistant Hypertension* has few retrieves (n=7). The retrieve count indicates how many distinct data requests are made, such as querying for all medications using codes from a specific value set, which can then be filtered or shaped in multiple ways.

### Expression Depths

Expression depth is an indicator of how many logical expressions are applicable concurrently, which is roughly correlated with how many phenotype definition criteria are concurrently applicable. Figure 8 provides a histogram of total expression depth as well as `where` clause expression depth. `where` clause expression depth is an indicator of how complicated data filtering expressions are. Finally, Figure 9 illustrates expression depth per expression category, which shows how many expressions of each different category are concurrently applicable.

**Figure 8.**
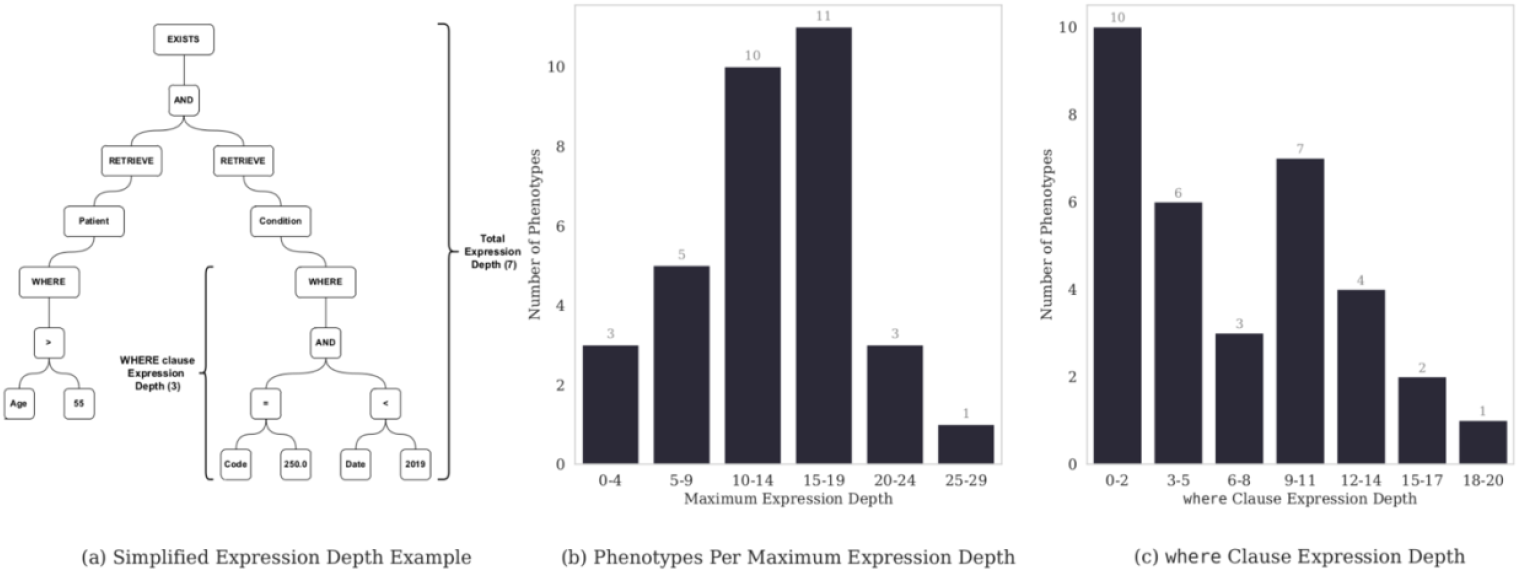
Example of expression depth (a), along with histograms of total (b) and `where` clause (c) expression depths.

**Figure 9.**
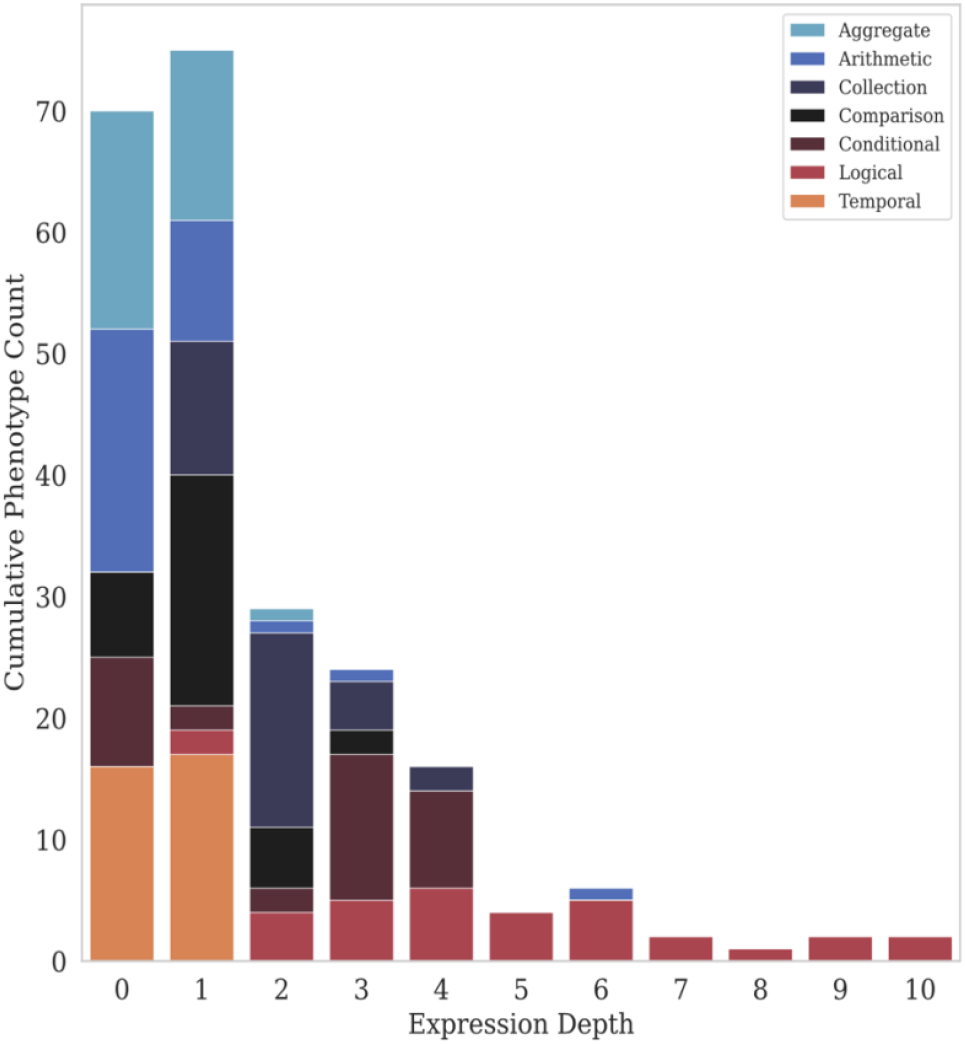
Expression depth utilization per category.

The lowest total expression depth is 4 (*Multiple Sclerosis, Crohn’s Disease*, and *Autism*) and the highest is 27 (*Familial Hypercholesterolemia*), with a median of 14 (mean: 13.5, std: 5.7). Seven phenotypes have a `where` clause expression depth of zero (*White Blood Cell Indices, Rheumatoid Arthritis, Multiple Sclerosis, Crohn’s Disease, Atrial Fibrillation, Autism*, and *Developmental Language Disorder*), meaning that data is only filtered by value set and no other criteria. *Red Blood Cell Indices* has the highest `where` clause expression depth of 18, and the median `where` clause expression depth is 7 (mean: 6.6, std: 5.4).

Many phenotypes have expression depths of zero or one for aggregate, arithmetic, collection, comparison, and conditional expression. Logical expressions always have a depth of at least one. There are some instances of expression depths in the two to four range for conditional, collection, comparison, and logical expressions, but only logical and arithmetic expressions have a depth of five or greater. Only logical expressions have a depth greater than six, with a maximum of 10 in two cases (*Red Blood Cell Indices* and *Familial Hypercholesterolemia*).

## Discussion

In this work we found that all of the 33 phenotypes evaluated use a combination of clinical logic and operational logic as part of their definitions. Similarly, despite the range of conditions, all of the definitions consisted of logical criteria, representing both clinical and operational logic, and tabular data, consisting of codes from standard terminologies and keywords for natural language processing. The original metadata associated with phenotype algorithms we evaluated was found to be irrelevant for this analysis or incorrect, demonstrating the need not only to identify more relevant metadata elements and ensure they are appropriately curated and verified, but also ensure metadata review and curation is ongoing to ensure accuracy. Some metadata elements that were originally curated by hand in PheKB, such as the categories of data or medical vocabularies used, could be extracted and updated programmatically if phenotype algorithms were represented in a standard computable format, such as CQL and FHIR. Overall, ongoing work is needed to improve metadata collection for phenotype algorithms, with some groundwork proposed in the field.[Chapman2021, Alper2022]

The artifacts that comprised the phenotype definitions can be divided into two high-level categories: logic and tabular data. The tabular data consists of value sets of codes from various code systems and lists of keywords or regular expressions used for NLP, although we did not analyze the latter in depth. We identified that diverse vocabularies are used for phenotype algorithms - all of which could be represented in CQL and FHIR. The OMOP common data model standardizes the terminologies used to a smaller subset, however, and comprehensive phenotyping platforms should anticipate accommodating a broad set of vocabularies.

Phenotype logic can be further divided into two categories: clinical logic and operational logic. Clinical logic is the core of the phenotype definition, and describes the clinical definition of the phenotype. Clinical logic includes things such as which diagnoses are relevant, which procedures, medications, and laboratory orders are associated with the phenotype, as well as patient demographic criteria that should be considered. Operational logic is also important, and while it contributes to the clinical definition, it is typically a bridge to how data are recorded in the EHR. Patterns in operational logic have been observed,[Rasmussen2014] but they can be difficult to express accurately using universally applicable logical expressions. For example, the *Herpes Zoster* phenotype requires that a matching patient have at least 5 years of continuous enrollment. The reason for this requirement is to “increase the probability that a subject’s status with respect to herpes zoster infection is known by the health care system.” This does not necessarily increase the correctness of the phenotype definition, but may nevertheless increase the negative predictive value. Another very common operational criterion is the requirement that a patient have at least 2 diagnoses of a given condition. This criterion is relatively simple to define using universally applicable CQL logic, while the concept of enrollment is determined differently at different institutions. One solution to this problem, which is available when using a modular formal representation, is to have local implementations for common operational criteria that are used during cohort execution. This is the same approach used in computer software, where system libraries provide routines with known names and well-defined parameters, but the implementation varies according to the operating system. However, this highlights the need for more broadly accepted metrics applicable to a wide range of health data that convey information about the completeness of capture of patient data, for a given data source. Completeness has been explored and reported in data quality frameworks,[Kahn2016, Schmidt2021] but work is needed to integrate these considerations more readily into phenotype authoring.

The types of expressions observed were simple, and required more complex calculations and aggregations of data to take place within the phenotype. The complexity of the phenotypes then comes from the topology-many simple expressions linked together in increasingly complex ways. The prevalence of certain expressions such as data retrieval, manipulation, and literal expressions is expected, but we were surprised by the relatively low usage of arithmetic and aggregate expressions, which indicates that in most cases data values are used directly, not used to construct derived values. Both the `count` and `sum` aggregate expressions are used as cardinality constraints (e.g., at least 2 diagnoses required) and not in an arithmetic context. The highly used existential operator (`exists`) is used for the same purpose (e.g., does an observation exist that meets certain requirements). Additionally, even though about two thirds of phenotypes use temporal expressions, the total number used is relatively small. The fact that the `and` operator is used about twice as much as the `or` and `not` operators makes sense, since conceptually, many phenotypes are defined by clusters of concurrent criteria rather than by the disjunction of many different criteria.

Overall expression depths seem to be normally distributed around 15, while `where` clause expression depth appears to be bimodal, but generally trends down at high values. The downward trend implies that complicated data filtering expressions are uncommon. Most expression categories are not very deeply nested, and only logical expressions (and in one case arithmetic expressions) have a depth of 5 or higher. This can be interpreted to mean that conceptually simple criteria are combined in complex ways using `and` and `or` expressions. This can be confirmed by looking at the flowcharts provided with some phenotype definitions, which may have a complicated topology, but the criteria represented by each node are relatively simple.

The total number of expressions per phenotype is generally not very high, with a median of just 59. The total number of expression types is also quite low, at just 43 (CQL has over 200 expression types). There are several possible reasons for the simplicity of the phenotypes in the data set. First, in our experience, implementing even simple phenotype definitions is quite challenging, so authors may choose to keep definitions simple to make implementation practical. Second, the phenotypes generally restrict themselves to data available in the EHR, which may be simplified, as this data is often for billing purposes. Further, since the PheKB phenotypes are designed to be shared, authors may limit themselves only to data available to most implementers, such as the most basic data elements. Finally, since phenotypes are created as narrative text, the lack of a formal expression language may be a factor that limits the level of detail provided.

There is no clear correlation between the severity or complexity of presentation of a disease and the number of expressions used. For example, something ostensibly simple like *Height* has ten times as many expressions as *Multiple Sclerosis*. There are also two Type 2 Diabetes phenotypes, one with 49 expressions and one with 100, so even the same disease can be represented in vastly different ways. This implies that a large part of phenotype definition complexity is determined by the level of detail that the author decided to use. This is independent of any formal representation, and may depend on the intended use of the phenotype definition.

### Limitations

We note the following limitations in this work. First, the data set used is relatively small, consisting of only 33 phenotype definitions, and there may exist additional phenotype definitions that would alter our conclusions. Additionally, many of the phenotypes evaluated were developed for conducting genomic studies, and the implementation decisions may have been tuned specifically for that purpose, biasing our findings. We also note that all the definitions in our dataset were designed to detect patients with a single condition. Even though EHR data provides the opportunity to conduct research on patients with multiple simultaneous conditions, the phenotype definitions we used were not designed to identify cohorts of such complex patients. The phenotypes analyzed are reminiscent of those that might be used for recruiting patients for randomized controlled trials, but we hope this work will provide some insights that lead to methods for developing higher fidelity phenotype definitions, and that these definitions can be used for so-called *deep* phenotyping.

The decision not to include NLP directives in our analysis was purposeful, but impacts any conclusions we would wish to draw about individual phenotypes. We know that most (n=28) phenotype definitions make use of some form of NLP, so we are omitting data from a significant number of definitions. However, almost all definitions including NLP directives simply provide a keyword list or regular expressions, and not higher-level entities, negation, and/or temporal constructs. In some cases, no details are given about how the NLP should be implemented. For example, the *Red Blood Cell Indices* phenotype definition says only “NLP was implemented to that regard” (referring to identifying patients taking specific medications). So, we believe that including a more detailed analysis of NLP data elements would not be informative due to their underspecificity in the dataset. A substantial amount of data useful for EHR-driven phenotyping may be stored in clinical notes. The body of research focused on extracting this data is growing,[Zeng2018] but to our knowledge, no widely accepted standard representation for NLP metadata and processes has yet emerged. The PhEMA research team is working on methods for integrating NLP into both FHIR [Hong2019] and CQL [Wen2021] and in future work we hope to integrate this research into our analysis of phenotype definitions using formal representations.

## Conclusion

In this work, we characterize a set of 33 validated research phenotype definitions, identifying how phenotype implementations use a combination of clinical logic and operational logic. The phenotypes analyzed are composed of the same high-level components, namely tabular data and logical expressions. The most important type of tabular data analyzed here are value sets, which can readily be represented in a standard format. Despite the limited number of expressions used from those available, individual phenotype algorithms could become complex in the total number of expressions needed and depth of nested operations, often to accommodate the needed operational bridge from how EHR data is collected and recorded. This complexity points to the need to use standard-based representations for expressing, sharing and implementation of phenotype definitions.

## Data Availability

All data produced in the present study are available upon reasonable request to the authors

## Acknowledgements

The authors wish to thank Frank Mentch from the Children’s Hospital of Philadelphia for providing feedback on an earlier draft of the manuscript.

## Footnotes

1 https://github.com/PheMA/phekb-phenotypes

2 https://github.com/PheMA/elm-utils

